# Development and Validation of an Evidence-based Home Pursed Lip Breathing Protocol for Improving Health Outcomes in Patients with COPD

**DOI:** 10.1101/2023.08.10.23293933

**Authors:** Houqiang Huang, Jun Da, Roger Watson, Mark Hayter, Min Huang

## Abstract

**Aim:** To develop and validate an evidence-based home PLB intervention protocol for improving related health outcomes (e.g. dyspnea, exercise capability, etc.) in COPD patients and to present a detailed intervention development process.

**Methods:** Phase one of the Medical Research Council Framework for Developing and Evaluating Complex Interventions was employed to guide the development process of the PLB intervention. The appropriateness of the PLB protocol was finally determined by a panel of experts using the content validity index.

**Results:** The preliminary PLB intervention protocol was developed based on several underlying rationales, which encompass the extension of expiration time, enhancement of respiratory muscle strength, augmentation of tidal volume, and integration of the most reliable research evidence obtained from four systematic reviews, six RCTs, five clinical trials, and ten recommendations. The PLB intervention was structured with a designated time ratio of inspiration to expiration, set at 1:2. Additionally, the training parameters of the PLB intervention were established as follows: three sessions per day, each lasting for 10 minutes, over a duration of 8 weeks. The intensity of PLB training was individualized, with the inhalation component adjusted according to each participant’s tolerance level, while emphasizing the exhalation phase to ensure complete expulsion of air from the lungs.

Regarding the content validity of the PLB intervention protocol, it demonstrated excellence, as consensus was achieved among all panel experts. Both the item-level and scale-level Content Validity Indices (CVIs) reached the maximum score of 1.0, indicating a high level of agreement and credibility in the content of the protocol as evaluated by the expert panel.

**Conclusion:** An optimal evidence-based home PLB protocol has been adapted and developed to manage COPD patients’ health-related outcomes. The protocol is transparent and fully supported by relevant mechanisms, concrete evidence, recommendations, and experts’ consensus.

**Impact:** The development of the PLB intervention protocol has shown considerable variation among studies, resulting in inconclusive effects in COPD patients. Moreover, there is currently a dearth of an evidence-based PLB intervention protocol, particularly in the home setting. Consequently, this study, guided by the MRC framework, has successfully formulated an evidence-based home PLB intervention aimed at enhancing the health outcomes of COPD patients

## 1 INTRODUCTION

Chronic Obstructive Pulmonary Disease (COPD) is a progressive respiratory condition characterized by persistent airflow limitation and represents a significant global health burden(Amir Sharafkhaneh, 2021). According to the World Health Organization, COPD is projected to become the third leading cause of death worldwide by 2030(Mathers & Loncar, 2006). Despite notable advancements in medical interventions, the home management of COPD remains challenging, necessitating the exploration of complementary therapeutic approaches to enhance health outcomes and improve patients’ quality of life.

Pulmonary rehabilitation has emerged as a fundamental aspect of COPD management, providing a comprehensive approach to enhance functional capacity, alleviate symptoms, and improve the overall quality of life for affected individuals(Spruit et al., 2013). Among the array of breathing exercises employed in pulmonary rehabilitation, pursed lip breathing (PLB) has garnered considerable attention owing to its potential to positively impact respiratory mechanics, increase oxygen saturation, and alleviate dyspnea(Weihua Zhang, 2018).

However, despite potential promising evidence regarding the benefits of PLB, notable gaps persist in the existing literature, leading to significant variations in the protocols utilized across different studies. First, the lack of clarity and specificity in existing PLB protocols makes it challenging for COPD patients to perform the exercises accurately without proper guidance. Second, the considerable variability in the duration, frequency, and intensity of PLB exercises among studies leaves healthcare providers with limited guidance when tailoring PLB interventions for individual patients(Holland et al., 2012; Roberts et al., 2013; Yang et al., 2022). Third, there is a significant gap in terms of a transparent and consensus-driven process for developing PLB intervention protocols in studies, which hinders the integration of PLB as a consistent and evidence-based therapeutic option in COPD management(Weihua Zhang, 2018). Last, most PLB research has been conducted in controlled clinical settings, and there is a paucity of evidence regarding the practicality and effectiveness of implementing PLB exercises in the home setting, where most COPD patients manage their condition daily(Weihua Zhang, 2018).

These identified gaps in the existing literature underscore the need for a comprehensive and validated evidence-based home PLB protocol that addresses the above limitations and offers clear guidelines for COPD patients to incorporate PLB exercises effectively into their self-management routine. The present research aims to bridge this critical gap by developing and validating an evidence-based home PLB exercise protocol designed for COPD patients. The MRC framework has been used to guide the process of the proposed protocol, which is grounded in current evidence from exercise mechanisms, systematic reviews, clinical trials, and recommendations(Skivington et al., 2021). By integrating existing evidence and tailoring interventions to address the unique challenges posed by COPD, the protocol seeks to optimize therapeutic outcomes and empower patients to play an active role in their self-management. Thus, the main objective of this study is to develop and validate an evidence-based home PLB protocol that can be further employed to improve health outcomes in COPD patients.

## 2 METHODS

### 2.1 Overview of this research design

This study is guided by the initial phase of the Medical Research Council (MRC) Framework for Developing and Evaluating Complex Interventions(Skivington et al., 2021). The primary objective is to develop a comprehensive and evidence-based home PLB intervention protocol specifically tailored for patients with COPD. To ensure a systematic and rigorous approach, the entire process encompasses three crucial and interconnected steps.

The initial step involves the justification for selecting PLB as the intervention and delving into the relevant mechanisms underlying its potential effectiveness in improving health outcomes for COPD patients. The second step centers on the meticulous identification and assessment of the most relevant and up-to-date evidence available. This entails an extensive review of the existing literature, including systematic reviews, clinical trials, RCTs, and recommendations related to PLB intervention in COPD. By synthesizing the available research evidence, the protocol can be developed by effective and evidence-based approaches. Finally, the third step focuses on the content validation of the developed PLB intervention protocol. To achieve this, a panel of experts specialized in respiratory-related professions is convened. These experts, with their wealth of experience in respiratory management, are essential in providing critical input and evaluating the proposed protocol’s validity.

By adhering to the Medical Research Council Framework and executing these three essential steps, this study aims to produce a robust, evidence-based, and carefully validated home PLB intervention protocol. This protocol, when implemented, has the potential to significantly contribute to the management and improvement of health outcomes for patients with COPD. Ethical approval for this study has been obtained from the Ethics Committee at the Affiliated Hospital of Southwest Medical University (KY2023105), China. The study design flowchart is illustrated in Figure 1. In addition, this study has been registered in the open science framework platform (https://doi.org/10.17605/OSF.IO/E4BXM).

### 2.2 Justification for PLB and identifying the relevant mechanisms

The justification for the use of PLB is of paramount importance in this study. Additionally, it is believed that the MRC framework emphasizes exploring underlying rationales to explain why an intervention may lead to the desired intervention effect^(Skivington^ ^et^ ^al., 2021)^. Prior to the development of the PLB intervention in this study, several databases and websites were searched to obtain relevant literature, including PubMed, the Cochrane Central Register of Controlled Trials (CENTRAL), Embase, JBI, Science Direct, the Chinese Biomedical Database (CBM), and Google Scholar, to justify the PLB and identify appropriate mechanisms. The search terms utilized included ’COPD’, ’chronic obstructive pulmonary disease’, ’breathing exercise’, ’pursed lip breathing’, ’mechanism’, and ’theory’. The search strategy was adjusted in each database accordingly. Currently, by gaining a thorough understanding of the physiological and therapeutic principles that support the use of PLB, the intervention protocol can be designed on a solid theoretical foundation and deepen an underlying understanding of using PLB as an effective intervention to improve exercise tolerance, and dyspnea in COPD patients.

### 2.3 Identification of Relevant Evidence

The MRC framework recommends that developing interventions systematically requires employing the best available evidence; thus, the first step is to search existing literature, including systematic reviews, clinical trials, and practice recommendations, to underpin the PLB protocol development by using optimal evidence. In this study, an extensive search was separately performed in PubMed, CENTRAL, Embase, JBI, Science Direct, CBM, and Google Scholar from their inception to July 2023 to identify current appropriate evidence associated with PLB intervention for improving health outcomes. Search terms such as ’COPD’; ’chronic obstructive pulmonary disease’; ’breathing exercise’; ’pursed lip breathing’; ’systematic review’; ’review’; ’meta-analysis’; ’recommendation’; ’guideline’; and ’trial’ were used for the electronic database search. Additionally, the reference lists of the literature were screened to identify related publications. In the end, research evidence for PLB protocol development was extracted from four systematic reviews(Holland et al., 2012; Roberts et al., 2013; Weihua Zhang, 2018; Yang et al., 2022), six RCTs(Bhatt et al., 2013; Elfa et al., 2019; Margaret A. Nield, 2007; Mohamed, 2019; Nield M A, 2007; Sakhaei et al., 2018), five clinical trials(Bianchi et al., 2007; Reid & Chung, 2013; Spahija et al., 2005; Van der Schans et al., 1995; Visser et al., 2011), and ten recommendations(Association, 2017; Clinic, 2017; Faling, 1986; Gronkiewicz, 2008; Illidi et al., 2023; K, 2017; Kumaran, 2018; Lareau SC, 2020; Marciniuk et al., 2011; Vatwani, 2019).

### 2.4 Content Validation of PLB Protocol

The MRC framework recommends that optimal complex interventions should be validated using the content validation approach, which consists of experts’ opinions and ratings of all the intended intervention components(Bleijenberg et al., 2018). Therefore, a six-step approach was undertaken to judge the content validation of the PLB, consisting of (1) formulation of the content validity assessment form, (2) selection of the review experts, (3) implementation of the content validation study, (4) review of domains and items, (5) rating each item with or without comments, and (6) estimation of the content validity(Yusoff, 2019).

The content validity assessment form consists of eight items, covering four aspects: justification of the intervention modality type, PLB techniques, PLB duration and frequency, and prepared conditions before PLB practice, respectively. Each item was independently rated by each enrolled expert using a 4-point Likert scale ranging from 1 to 4, where ’1’ indicates ’totally inappropriate,’ and ’4’ indicates ’very appropriate.’ Each item rated with a score lower than 3 by any panel expert is required to give feedback and suggestions with concrete evidence to further help refine the PLB intervention protocol. For validation experts’ recruitment, a purposive sampling method was adopted through the research team’s network. The inclusion criteria for experts were: (1) clinical or research experience in respiratory management for at least 10 years; (2) assistant professors or above; and (3) having a master’s degree or above. According to a suggested study(Lynn, 1986), each item should achieve an item-level Content Validity Index (CVI) value of 0.83. Therefore, a panel of six experts is an appropriate quantity for this purpose. The scale-level CVI was determined to be satisfactory for PLB if the proportion of items scored as ≥3 was ≥80%(Lynn, 1986). If the CVIs could not reach satisfactory values in one round, the PLB protocol would be further refined based on the suggestions received from the panel members. Rounds of assessment would be repeated until the pre-set CVI values were achieved.

## 3 RESULTS

### 3.1 The PLB intervention protocol

The content of PLB intervention protocol includes the following components: (1) justification of PLB modality type, (2) identification of PLB techniques, (3) identification of PLB duration and frequency, and (4) prepared conditions before PLB practice. Details of the PLB intervention protocol, along with relevant supporting evidence, are illustrated in Table 1 below.

### 3.2 Justification of PLB Modality Type

The PLB was underpinned by evidence derived from two systematic reviews(Weihua Zhang, 2018; Yang et al., 2022), three recommendations(Illidi et al., 2023; Lareau SC, 2020; Marciniuk et al., 2011), four RCTs studies(Bhatt et al., 2013; Elfa et al., 2019; Margaret A. Nield, 2007; Sakhaei et al., 2018), and two clinical trials(Bianchi et al., 2007; Spahija et al., 2005), as well as likely effective intervention mechanisms of PLB. In this study, PLB is proposed as a breathing exercise based on current evidence suggesting its likely effectiveness and its inexpensive, non-invasive, and non-pharmacological characteristics(Marciniuk et al., 2011; Sakhaei et al., 2018; Weihua Zhang, 2018; Yang et al., 2022), making it a cost-effective intervention for COPD patients with poor economic conditions. Additionally, this training method is simple and easy to learn, without the restrictions of equipment and location(Li et al., 2021). It is highly favored by older individuals, such as COPD patients, and has been demonstrated to reduce interference with daily physical activities(Li et al., 2021). Furthermore, COPD patients can experience more significant benefits from conducting PLB at home compared to other types of breathing exercises(Illidi et al., 2023; Sakhaei et al., 2018), although a standardized home-based PLB protocol is still lacking(Weihua Zhang, 2018). Therefore, PLB has been recommended by the Canadian Thoracic Society and the American Thoracic Society to manage COPD patients, respectively(Lareau SC, 2020; Marciniuk et al., 2011).

As for the underlying rationales, it is believed that the mechanism by which PLB exerts improved exercise tolerance is by prolonging expiration time during PLB exercises to help prevent airway collapse and reduce pulmonary hyperinflation(Bhatt et al., 2013; Bianchi et al., 2007; Illidi et al., 2023; Spahija et al., 2005). This effect leads to better ventilation and gas exchange, allowing patients to sustain physical activity for longer durations. Additionally, the selection of PLB for relieving dyspnea is attributed to sustained PLB training leading to an increase in respiratory muscle strength(Margaret A. Nield, 2007). With greater respiratory muscle strength, less force is generated with each breath, which may reduce motor input and output to the respiratory muscles, ultimately decreasing the perceived sense of respiratory effort. Furthermore, PLB can contribute to an increase in the quality of life for COPD patients by enhancing respiratory function. Specifically, the activation of abdominal muscles during expiration helps increase tidal volume, improving gas exchange and oxygen saturation(Elfa et al., 2019; Illidi et al., 2023). Additionally, the reduction in chest wall volume at end-expiration alleviates dyspnea, anxiety, and tension, ultimately leading to an improved sense of well-being and overall quality of life for individuals with COPD(Bianchi et al., 2007; Elfa et al., 2019).

### 3.3 Identification of PLB Techniques

The PLB techniques for this study were identified and adapted through concrete evidence from seven recommendations(Association, 2017; Clinic, 2017; Faling, 1986; Gronkiewicz, 2008; K, 2017; Kumaran, 2018; Vatwani, 2019), and one clinical trial(Van der Schans et al., 1995).

Traditionally, PLB generally requires inhaling through the nose for at least 2 seconds with the mouth closed and then slowly exhaling all the air in the lungs with lips pursed, similar to whistling or gently flickering the flame of a candle, for at least 4 seconds(Clinic, 2017; Vatwani, 2019). However, this modality may not perfectly meet the requirements of COPD patients training at home due to inexplicit training instructions, uncertain training time, intensity, etc. As is known, the core point of PLB is to obtain a longer expiration than inspiration with an appropriate time ratio between them. Based on current evidence involving COPD, it is more likely recommended to support that the exhalation time is twice that of the inhalation time(Association, 2017; K, 2017; Van der Schans et al., 1995). In practice, the current evidence indicates that it is beneficial to deliver careful and explicit repeated training instructions to COPD patients ^(Gronkiewicz, 2008; K, 2017; Vatwani, 2019)^ and let patients count during PLB training(Vatwani, 2019), so that the explicit time of the inspiration and expiration for PLB in this field should be stated to guide the process of training, which would complement the existing PLB modality to better fit COPD patients training at home.

Therefore, the expiration time determined to be two times the inspiration time is restricted to four seconds in this study as it is the minimum duration in one required exhalation and determined by a consensus of the research team. As for the intensity, existing evidence was in favor of the depth of inhalation relying on individual tolerance (Gronkiewicz, 2008), but exhalation should involve expelling all air from the lungs(Clinic, 2017). To achieve the optimal outcome, air must be breathed into the lungs through the nostrils in this technique, considering that air can be heated, purified, moisturized, and pressurized(Faling, 1986; Kumaran, 2018).

### 3.4 Identification of PLB Duration, Frequency, and Intensity

PLB duration and frequency were determined based on evidence from three systematic reviews(Holland et al., 2012; Roberts et al., 2013; Yang et al., 2022). Existing systematic reviews reported a wide variation in PLB duration, ranging from a minimum of 5 minutes per session to 60 minutes per day in studies that mentioned training time. Additionally, the frequency of PLB ranged from twice daily to five times a day, with durations varying from 4 weeks to 6 months(Holland et al., 2012; Roberts et al., 2013; Yang et al., 2022). Given the potential limited physical tolerance of COPD patients, it is essential to consider a tolerable and effective training time to avoid discomfort during home-based PLB training while developing the PLB protocol.

The current evidence supports the feasibility of a three-times-daily PLB intervention as the most common frequency, with 10 minutes per session being considered acceptable(Holland et al., 2012; Roberts et al., 2013; Yang et al., 2022). Considering both feasibility and COPD patients’ tolerance, these three daily sessions should be practiced separately in the morning, at noon, and in the evening. This adaptation is supported by current research evidence and the expertise of the research team, specialized in chronic respiratory disease management. Furthermore, the PLB intervention duration was specifically set for 8 weeks, based on evidence from the Cochrane systematic review, which demonstrated definite and significant effects on dyspnea and exercise tolerance(Holland et al., 2012).

### 3.5 Prepared Conditions before PLB Practice

Practicing PLB, a COPD patient should be in a comfortable and safe environment(Association, 2017; Reid & Chung, 2013), maintaining an upright, slightly forward posture, which can have beneficial effects on improving respiratory rate and reducing breathlessness during training^(Mohamed, 2019)^. Additionally, patients are advised to relax the muscles of the shoulders and neck before practice(K, 2017; Sakhaei et al., 2018). Furthermore, it is recommended to have a chair or other leaning object nearby during the training, which COPD patients can hold onto to avoid potential risks such as falls (Nield M A, 2007; Visser et al., 2011).

### 3.6 Validation of the PLB Intervention Protocol

A total of six experts specialized in respiratory-related professions were invited to participate in this validation study of the PLB intervention protocol. All the included experts had more than 10 years of professional experience in respiratory management, with one-third of them having a doctorate degree and the rest possessing a master’s degree. The panel of experts consisted of two clinical physicians, two clinical nurses, and two university researchers, all holding at least an assistant senior professional qualification. The detailed characteristics of the experts are listed in Table 2. The content validity consensus was obtained through one round of assessment, in which each item of the PLB intervention protocol was rated by the six experts with a score of ’3’ (appropriate) or ’4’ (very appropriate). Additionally, one expert suggested a revision to the training time, reducing it from 10 to 5 minutes. In this validation study, the CVIs at the item-level and scale-level had reached an ideal score of 1.0. Detailed results of the experts’ assessments are presented in Table 3.

## 4 DISCUSSION

### 4.1 The successful development of the PLB intervention protocol

This study ultimately developed an evidence-based PLB intervention protocol aimed at improving the health status of COPD patients who can practice at home. The unique aspect of this study is that the PLB intervention was formulated based on the MRC framework.

The MRC framework emphasizes the importance of evidence in supporting intervention development(Skivington et al., 2021). For this study, evidence from systematic reviews(Holland et al., 2012; Weihua Zhang, 2018; Yang et al., 2022), RCTs (Bhatt et al., 2013; Elfa et al., 2019; Margaret A. Nield, 2007; Mohamed, 2019; Nield M A, 2007; Sakhaei et al., 2018), clinical trials(Bianchi et al., 2007; Reid & Chung, 2013; Spahija et al., 2005; Van der Schans et al., 1995; Visser et al., 2011), and recommendations(Association, 2017; Clinic, 2017; Faling, 1986; Gronkiewicz, 2008; Illidi et al., 2023; K, 2017; Kumaran, 2018; Lareau SC, 2020; Marciniuk et al., 2011; Vatwani, 2019) served as a crucial foundation in theorizing the PLB, selecting and adapting the appropriate modality, techniques, duration, frequency, and preparation before training. Additionally, the PLB protocol took into careful consideration the specific characteristics, physical tolerance, and training-supporting environment of COPD patients, ensuring that the frequency and duration of the PLB were sufficient to achieve the intended health outcomes while also being practically feasible.

Several mechanisms were identified to understand how the PLB can contribute to the expected results. PLB, through intentional prolongation of expiration time, increasing respiratory muscle strength, raising tidal volume, and reducing chest wall volume, can offer significant benefits for COPD patients. The intentional prolongation of expiration time during PLB exercises is particularly valuable for COPD patients as it facilitates complete emptying of the lungs, preventing air trapping and reducing hyperinflation ^(Bhatt et al., 2013; Bianchi et al., 2007; Illidi et al., 2023; Spahija et al., 2005)^. This, in turn, alleviates the sensation of breathlessness and improves breathing efficiency. By engaging in prolonged exhalation, COPD patients enhance their ability to expel trapped air and stale gases, leading to improved gas exchange and better blood oxygenation.

Moreover, PLB exercises play an instrumental role in strengthening the respiratory muscles. As COPD often leads to respiratory muscle weakness due to the increased effort required to breathe against obstructed airways, strengthening these muscles becomes crucial for improved respiratory function. Regular practice of PLB can help increase the strength and endurance of the diaphragm and intercostal muscles, both of which play pivotal roles in the breathing process(Margaret A. Nield, 2007).

Furthermore, PLB contributes to an increase in tidal volume, which refers to the amount of air moved in and out of the lungs during a single breath^(Elfa^ ^et^ ^al., 2019; Illidi^ ^et^ ^al., 2023)^. By focusing on slow, deep breathing, COPD patients can maximize their tidal volume, allowing more air to reach the deeper regions of the lungs where gas exchange occurs. This, in turn, helps to enhance lung ventilation and optimize oxygen uptake while facilitating the removal of carbon dioxide from the body.

Additionally, PLB can help reduce chest wall volume in COPD patients^(Bianchi^ ^et^ ^al., 2007; Illidi^ ^et^ ^al., 2023).^. Over time, COPD can cause changes in the structure and flexibility of the chest wall, leading to increased chest expansion and reduced respiratory efficiency. However, by promoting relaxation of the shoulder and neck muscles and encouraging the use of proper posture during PLB practice, COPD patients can minimize excessive chest wall movement and create a more efficient breathing pattern.

Therefore, this breathing exercise intervention has a solid theoretical foundation to support its widespread use.

In addition, the content validation assessment for this PLB protocol yielded excellent results, with satisfactory scores achieved on both the item-level and scale-level of CVI after a single round of investigation involving feedback from the panel experts. However, one expert from the panel suggested shortening the session time from 10 to 5 minutes, as they considered it to be a little long and potentially difficult for COPD patients to accomplish. After thorough discussion among the research team and a review of existing evidence, no further revision was made. This decision was based on the current evidence, which indicates the feasibility of adopting a 10-minute intervention time(Roberts et al., 2013; Yang et al., 2022). Moreover, longer training time has been found beneficial for COPD patients, enabling them to gain sufficient practice and develop expertise and knowledge of PLB, ultimately leading to better outcomes. As a result, this protocol has been effectively validated.

### 4.2 Limitations

This study has several limitations. COPD patients, as the target users of this PLB intervention protocol, were not directly involved in the process of intervention development. However, conducting interviews with COPD patients before practicing the PLB intervention might have been challenging, as they may lack prior experience with PLB and may not be able to provide a proper assessment of the intervention protocol. Nevertheless, it is important to note that this intervention protocol development was only the first phase of a multi-phase research project. In the next stage of the project, COPD patients will be actively engaged to develop a digital theory-based pursed lip breathing intervention that considers their perspectives and specific needs. This will be followed by a rigorous pilot study design to assess the feasibility, acceptability, and preliminary effects of the intervention.

## 5 CONCLUSION

In conclusion, this study adopted the MRC framework and successfully developed an evidence-based home PLB intervention protocol to improve health conditions in patients with COPD. The development of the PLB intervention protocol was based on relevant mechanisms, systematic reviews, clinical trials, recommendations, and experts’ consensus. The established PLB intervention protocol will be further utilized to create a digital theory-based PLB intervention. To assess its feasibility, acceptability, and preliminary effects on health outcomes in COPD patients, a pilot study design and a qualitative study will be conducted. This rigorously designed PLB intervention protocol serves as an exemplary demonstration of developing a healthcare intervention using the MRC framework, offering valuable guidance to other researchers seeking to inform the development of their own evidence-based complex interventions.

## Data Availability

All data produced in the present work are contained in the manuscript

## Acknowledgements

This paper has not been published elsewhere in whole or in part. All authors have read and approved the content, and agree to submit it for consideration for publication in your journal. There are no ethical/legal conflicts involved in the article.

