## Supplementary material for "Development and Validation of an Evidence-based Home Pursed Lip Breathing Protocol for Improving Health Outcomes in Patients with COPD": Tables

**Table 1 Details of the PLB Intervention Protocol supported by relevant evidence**

| **Procedure** | **Content** | **Justification** |
| --- | --- | --- |
| Intervention modality type | Pursed lip breathing | - By prolonging expiration time to prevent airway collapse and then reduce pulmonary hyperinflation, exercise tolerance is improved(Bhatt et al., 2013; Bianchi et al., 2007; Illidi et al., 2023; Spahija et al., 2005) - A sustained increase in inspiratory and expiratory muscle strength for relieving dyspnea(Margaret A. Nield, 2007) - PLB may improve the quality of life by increasing the tidal volume through activating abdominal muscles during expiration (Elfa et al., 2019; Illidi et al., 2023) and reducing the chest wall volume at end-expiration(Bianchi et al., 2007; Illidi et al., 2023). - An easy, inexpensive, non-invasive, non-pharmacological breathing exercise intervention (Sakhaei et al., 2018; Yang et al., 2022) - PLB is likely to be effective, and a standardized PLB protocol is recommended to be developed in the home setting (Marciniuk et al., 2011; Weihua Zhang, 2018) - PLB seems to have more benefits than other types of breathing exercises(Illidi et al., 2023) - COPD patients at home can have beneficial effects from the PLB(Sakhaei et al., 2018) - PLB has been advocated by the Canadian Thoracic Society and the American Thoracic Society(Lareau SC, 2020; Marciniuk et al., 2011) |
| PLB  techniques | Breathe in through nose for 2 seconds<Count to 2> with mouth closed, and the depth of inhalation depends on the patient's tolerance | - Air inhaled through nose can be heated, purified, moisturized, and pressurized(Faling, 1986; Kumaran, 2018). - Careful and explicit repeated training instructions are beneficial, and the more frequently recommended inhalation time through nose is 2 seconds, which is counted by the patient’s own during training (Gronkiewicz, 2008; K, 2017; Vatwani, 2019) - The ability to tolerate inhalation depth is distinct among different individuals (Gronkiewicz, 2008) |
|  | Pucker your lips and exhale slowly all the air in the lungs through the small gap between lips for 4 seconds<Count to 4> | - It is recommended that expected exhalation time is 2 times than that of inhalation time(Association, 2017; K, 2017; Van der Schans et al., 1995) - Counting by patient’s own to calculate time of breathing out is helpful(Vatwani, 2019) - Recommend to slowly exhale all the air in patient’s lungs with lips pursed like whistling or gently flickering the flame of a candle”(Clinic, 2017) |
| Duration and frequency | 3 times a day, once in the morning, once at noon, and once in the evening | - PLB training with 3 times daily is feasible(Holland et al., 2012; Roberts et al., 2013) - The most frequently adopted frequency of PLB in clinical studies(Roberts et al., 2013; Yang et al., 2022) |
|  | 10 minutes per time | - 10 minutes of each training is feasible, and most studies have applied this time (Roberts et al., 2013; Yang et al., 2022) |
|  | For 8 weeks | - 8 weeks of PLB training showed definite and significant effects on dyspnea and exercise tolerance(Holland et al., 2012) - Often adopted duration in clinical studies(Roberts et al., 2013; Yang et al., 2022) |
| Prepared conditions before PLB practice | Sit or stand in a comfortable and safe environment and maintain an upright, slightly forward posture, and have a chair or other leaning objects nearby for patients in case of an emergency during the training in that case they can have support | - It is recommended to perform PLB in an upright and slightly forward posture in a comfortable and safe environment (Association, 2017; Reid & Chung, 2013) - Slightly forward leaning positioning is beneficial for improving respiratory rate and reducing breathlessness(Mohamed, 2019) - Chair or other leaning objects nearby are helpful to avoid potential risks during the training at home(Nield M A, 2007; Visser et al., 2011) |
|  | Relax the muscles of the shoulders and neck | - Recommended behaviors before performing PLB (K, 2017; Sakhaei et al., 2018) |

**Table 2 Characteristics of the panel expert**

| Expert characteristics | Number of Experts | Percentage (%) |
| --- | --- | --- |
| Sex |  |  |
| Male | 2 | 33.3% |
| Female | 4 | 66.7% |
| Country |  |  |
| China | 3 | 50.0% |
| Australia | 2 | 33.3% |
| America | 1 | 16.7% |
| Age |  |  |
| 30-40 y | 1 | 16.7% |
| 40-50 y | 2 | 33.3% |
| 50-60 y | 3 | 50.0% |
| Profession |  |  |
| Medicine | 4 | 66.7% |
| Nursing | 2 | 33.3% |
| Institution |  |  |
| University | 2 | 33.3% |
| Hospital | 4 | 66.7% |
| Working Position |  |  |
| Medical consultant | 2 | 33.3% |
| Nursing consultant | 2 | 33.3% |
| Researcher | 2 | 33.3% |
| Highest academic qualification |  |  |
| Doctorate degree | 2 | 33.3% |
| Master’s degree | 4 | 66.7% |
| Professional qualification |  |  |
| Senior physician | 2 | 33.3% |
| Senior nurse | 1 | 16.6% |
| Assistant senior nurse | 1 | 16.6% |
| Professor | 1 | 16.6% |
| Assistant professor | 1 | 16.6% |
| Years of professional experience |  |  |
| 10 years ~20 years | 3 | 50% |
| >20 years | 3 | 50% |

**Table 3 Content Validity of the Home PLB Intervention Protocol**

| Item | Description of Item | Results of Content Validity Assessment(experts number n=6) | | | |
| --- | --- | --- | --- | --- | --- |
|  |  | Number of experts rating "very appropriate" (4) | Number of experts rating "appropriate" (3) | Total number of experts rating content valid (3 or 4) | CVI |
| 1 | Pursed lip breathing | 6 | 0 | 6 | [Item-level] 1.00 |
| 2 | Sit in a comfortable and safe environment and maintain an upright posture with slightly forward posture, and have a chair or other items nearby for patients in case of an emergency during the training in that case they can have support | 5 | 1 | 6 | [Item-level] 1.00 |
| 3 | Relax the muscles of the shoulders and neck | 6 | 0 | 6 | [Item-level] 1.00 |
| 4 | Breathing in through nose for 2 seconds<Count to 2>with mouth closed, the depth of inhalation depends on the patient's tolerance. | 6 | 0 | 6 | [Item-level] 1.00 |
| 5 | Pucker your lips and exhale slowly all the air in the lung through the small gap between lips for 4 seconds<Count to 4> | 5 | 1 | 6 | [Item-level] 1.00 |
| 6 | Duration for 8 weeks | 5 | 1 | 6 | [Item-level] 1.00 |
| 7 | Practice 3 times a day, once in the morning, once at noon, and once in the evening | 4 | 2 | 6 | [Item-level] 1.00 |
| 8 | Practice 10 minutes per time | 2 | 4 | 6 | [Item-level] 1.00 |
| Scale-Level CVI of the whole BE protocol | |  |  |  | [Scale-level] 1.00 |

PLB=pursed lip breathing, CVI=content validity index
