## Supplementary figures and images for "Development and Validation of an Evidence-based Home Pursed Lip Breathing Protocol for Improving Health Outcomes in Patients with COPD"

### Figure 1

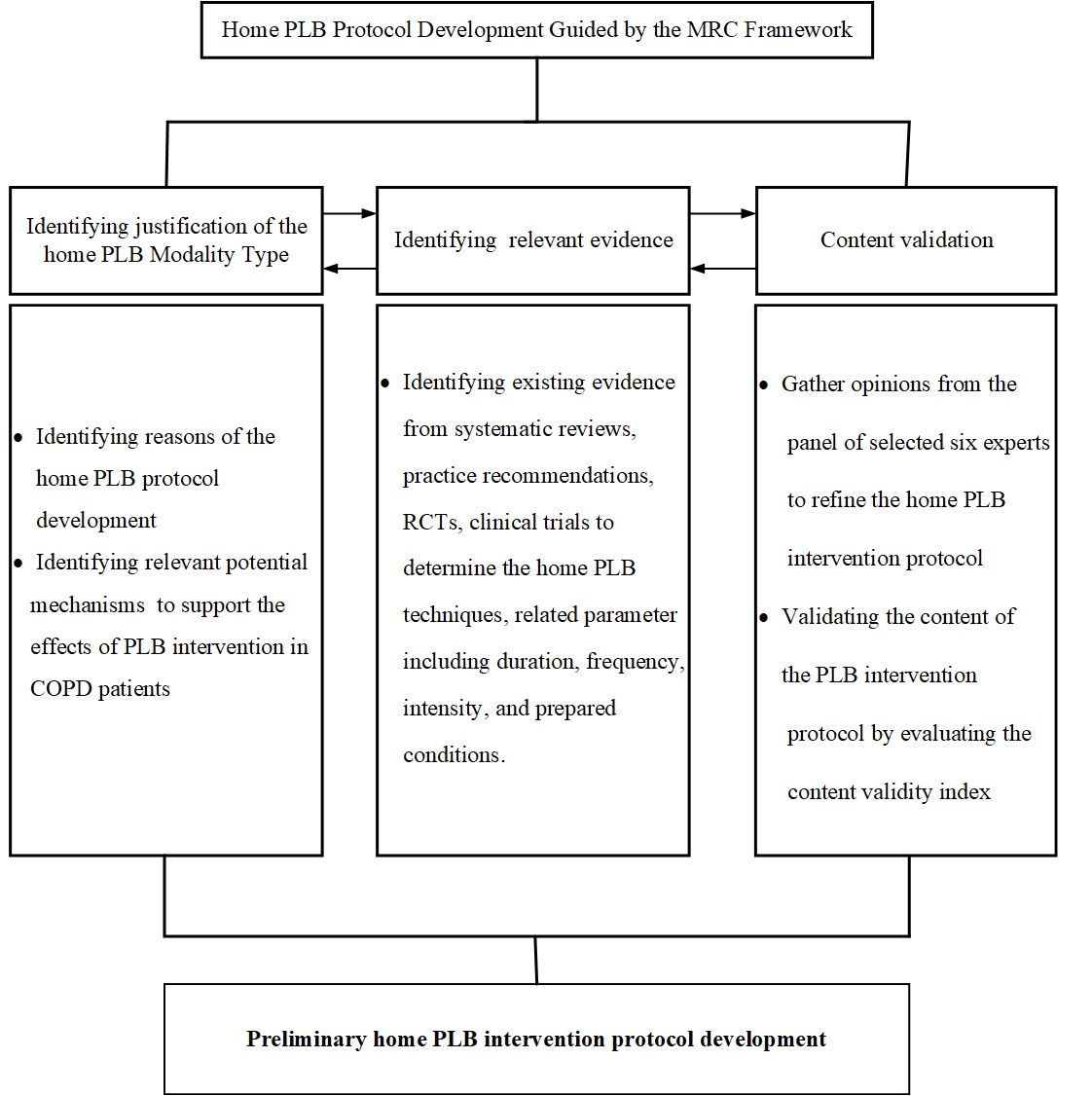
